# Do self-help groups improve sexual and reproductive health and HIV outcomes among female sex workers in sub-Saharan Africa? A scoping review

**DOI:** 10.1101/2024.10.10.24315253

**Authors:** Gracious Madimutsa, Fortunate Machingura, Owen Nyamwanza, Frances M Cowan, Webster Mavhu

## Abstract

**Introduction:** Self-help groups (SHGs) have been effective in improving the health and wellbeing of women generally but there is little evidence on whether and how they improve HIV and sexual and reproductive health (SRH) outcomes among female sex workers (FSWs), particularly in sub-Saharan Africa. This scoping review seeks to address this gap by identifying and analysing literature on SHG for FSWs in sub-Saharan Africa.

**Materials and methods:** This scoping review (1) identified relevant studies; (2) selected the studies; (3) charted the data; and (4) collated, summarised, and reported the results. A search strategy was developed; CINAHL, Medline and Global Health databases were searched.

**Results:** Eleven studies were identified, two were quantitative, seven were qualitative and two were mixed methods. Studies were from seven countries in sub-Saharan Africa. The studies suggested that SHGs can improve SRH outcomes and reduce HIV vulnerabilities among FSWs by providing emotional and financial support, health education, linkage to care, and social capital (i.e., benefits derived from associations). The studies also highlighted the need for tailored interventions that address the unique needs and challenges faced by FSWs.

**Conclusions:** The findings of this scoping review underscore the importance of building social cohesion by incorporating SHGs into a range of HIV prevention strategies in sub-Saharan Africa. SHGs have the potential to improve SRH and HIV outcomes among FSWs. Further research is needed to explore the effectiveness of SHGs in different contexts and to identify best practices for implementing and sustaining SHGs for FSWs.

## Introduction

Globally, female sex workers (FSWs) are 30 times more likely to acquire HIV than general population women [1]. High numbers of sexual partners, inability to control prevention measures [2], stigma that fuels violence and, criminalisation of sex work, combine to reduce FSWs’ exposure to services and support, which in turn, increases their vulnerabilities to HIV and other sexually transmitted infections (STIs) [3]. A systematic review of community empowerment and involvement of FSWs in targeted sexual and reproductive health (SRH) interventions in Africa found that FSWs aged 15 to 49 years were at a higher risk of SRH morbidity, violence and discrimination compared to same age general population women [4]. However, the overall impact of empowerment interventions on FSWs was not clearly established in this review.

In sub-Saharan Africa (SSA), the epicentre of the HIV epidemic [5] the burden of HIV and other STIs among FSWs is disproportionately high [6]. For example, in Zimbabwe, HIV prevalence was estimated to be 48% among FSWs in 2022 compared to 11% among the general population of adult women [7]. It is important to note that sex work has been a contributing factor in the spread of HIV in SSA [8].

FSWs’ vulnerabilities complicate their ability to prevent or manage HIV; therefore, interventions that address these vulnerabilities are needed. Self-help groups (SHGs), where individuals with commonalities come together to support each other, have potential for impact [9]. The concept of the SHG as a catalyst for change came about in 1935 when these groups were used to help alcoholics recover [10]. The approach became widely accepted for non-alcohol addiction problems after World War II [11]. In the 1960s, civil rights movements began to evolve in many developed countries, as people became aware of their collective power. This concept emphasised high levels of group ownership, control and management concerning goals, processes and outcomes [4]. According to Brody et al [12], collective or individual empowerment can happen when individuals join together to address their challenges.

SHGs may facilitate empowerment and foster autonomy in individuals or groups. Among the theories that explain how SHGs influence empowerment is the integrated empowerment framework for FSWs, which combines various elements and strategies aimed at empowering individuals or communities [13]. It integrates multiple aspects of empowerment and shows how they might achieve meaningful and sustainable outcomes [13].

Past research has shown that SHGs, along with other community mobilisation and structural interventions, can empower FSWs to address their specific vulnerabilities [14]. A study in India highlighted the impact of SHGs among sex workers, when FSWs who attended SHGs demonstrated higher HIV knowledge, accessed services more frequently, and were more likely to turn away clients who refused to use condoms, compared to those that did not attend SHGs [15]. However, there is a lack of substantive evidence on how SHGs can address health-related outcomes of FSW in the SSA context. This scoping review sought to explore whether and how SHGs improve FSWs’ SRH and HIV outcomes in SSA.

## Methods

The main objective of the scoping review was to explore a body of literature to identify what is known about SHGs in relation to addressing FSWs’ SRH and HIV outcomes in SSA. A preliminary search of MEDLINE, the Cochrane Database of Systematic Reviews and *JBI Evidence Synthesis* was conducted and no current or planned systematic or scoping reviews focusing on FSWs on the topic were identified. The scoping review started with the development of a study protocol. The study protocol was uploaded on Open Science Framework (OSF) and subsequently published [16].

### Identifying the research question

#### Review question

*The scoping review sought to answer the following questions:*

1. Do self-help groups improve SRH and HIV outcomes such as family planning and condom use, HIV/STI knowledge and prevention, HIV testing, treatment, and adherence, sexual self-efficacy, gender-based violence education, SRH knowledge/education, among female sex workers in SSA?

a. If so, what are the mechanisms through which SHGs improve these outcomes?
b. How feasible and acceptable are SHGs among FSWs?
c. What gaps exist in the evaluation of SHGs and SRH and HIV outcomes among FSWs in SSA?

### Eligibility criteria

The Population–Concept–Context (PCC) framework [17] was used to develop the research objective(s) and question(s) to inform inclusion and exclusion criteria and consequently, the literature search strategy.

#### Population

Female sex workers in SSA were included. The review considered FSWs as women who received money and/or goods or favours in exchange for sex. Transactional sex relationships were also considered as sex work, even if participants did not self-identify as sex workers. The review was restricted to studies conducted within SSA between 1 January 2000 and 30 September 2024. This period was chosen because it allowed the search to include relevant articles within the SSA context considering SHGs are relatively new within this setting compared to others such as South Asia where they have a longer and institutionalised history [18]. This period also considered that there was no similar scoping review during that time.

#### Concept

The scoping review explored the concepts of SHGs and FSWs’ SRH and HIV outcomes.

#### Context

Research studies conducted in sub-Saharan Africa.

### Types of Sources

The scoping review considered studies that met the inclusion criteria. Extracted quantitative data included characteristics of the study population, the proportion of the study population that consisted of FSWs, sample size, year of data collection, study location, sampling strategy and participants’ age range. The review also considered qualitative studies that reported on grounded theory, ethnography, case studies and qualitative descriptions.

### Study design

The framework developed by Arksey and O’Malley [19], which Levac et al. [20] and Colquhoun et al.[21] expanded on, and Peters [22] further outlined in the Joanna Briggs Institute Manual (2020 version) was employed. The stages of the review included: 1) identifying the research question, 2) identifying relevant studies, 3) selecting studies, 4) charting the data, and 5) collating, summarising, and reporting the results [19].

#### Stage 1: Identifying the research question

The Population–Concept–Context (PCC) framework [17] was used to identify the main concepts in the primary review question and to inform the search strategy. The primary review question was, ‘Do self-help groups improve sexual and reproductive health and HIV outcomes among female sex workers in sub-Saharan Africa?’. A sub-question to this was ‘If so, what are the mechanisms through which SHGs improve these outcomes?’ and another sub-question was ‘How feasible and acceptable are SHGs among FSWs?’. Yet another sub-question was ‘What gaps exist in the evaluation of SHGs and SRH and HIV vulnerabilities among FSWs in SSA?’. These questions enabled us to map the range of relevant literature around these aspects and inform the direction of future research.

#### Stage 2: Identifying relevant studies

##### Search strategy and information sources

Identification of studies relevant to this review was achieved by searching electronic databases of the published literature including Medline, Global Health and CINAHL databases. The general search strategy is outlined in Table 1:

**Table 1:**
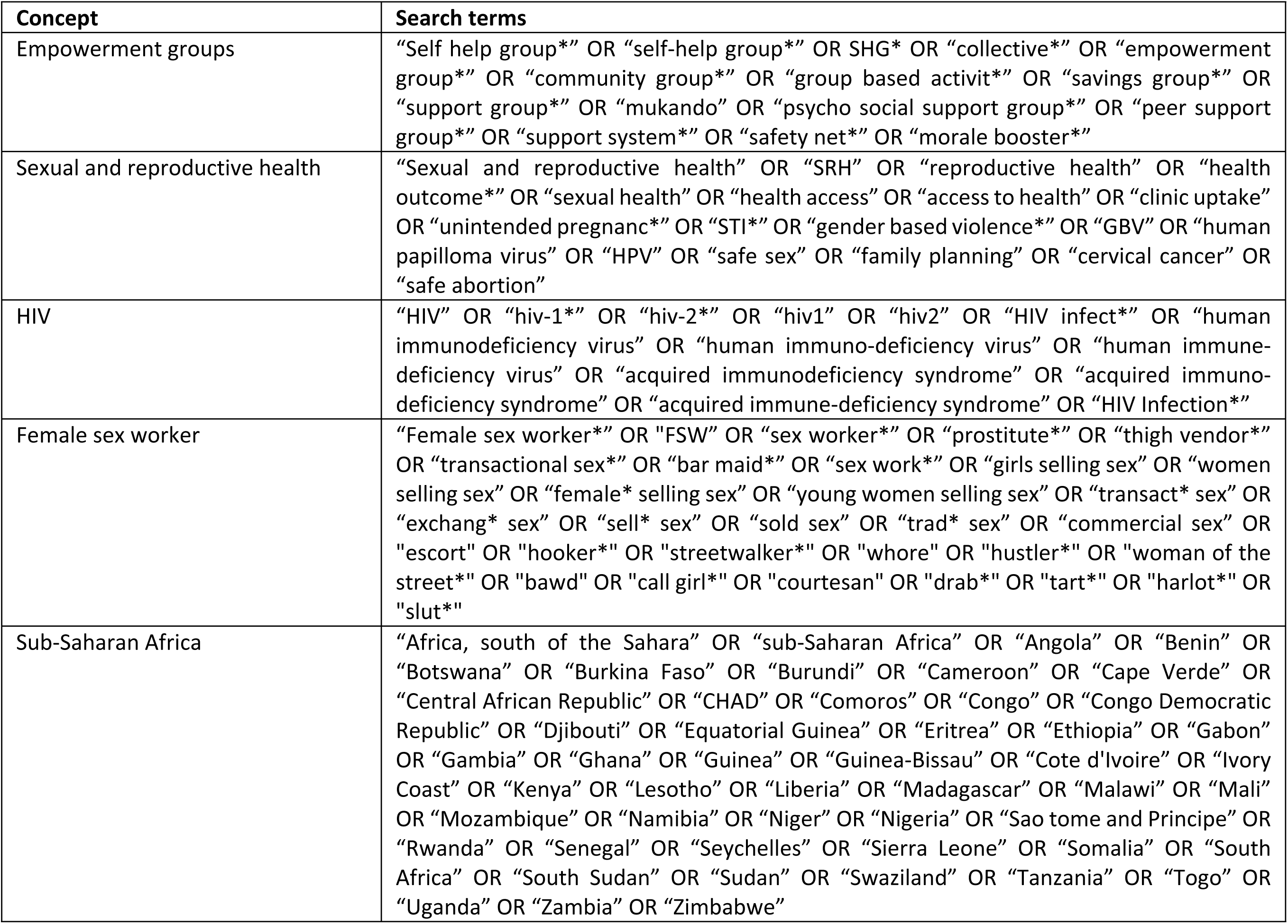
Search Strategy.

On 20^th^ April 2024, the lead author (GM) tested a general search strategy by running it using a Boolean search on the Discover platform, combining the search terms from the PCC (Population, Concept Context) framework using “AND” and separating different terms using “OR”. She then searched each of three databases (Medline, Global Health and CINAHL) using the key terms from the PCC framework to find the Medical Subject Headings (MeSH) used in the databases. She then combined the MeSH terms with the free text in the search strategy that she developed (see S1 Table). These databases were carefully selected for their comprehensiveness in covering the area under research. The researchers’ familiarity with the databases also enhanced efficiency in the search process. Search terms were determined with input from the research team, research collaborators and knowledge users. Searches were limited to literature published in English only and literature published before 2000 was excluded so that the review considered the most recent publications. Search results were downloaded and imported into Endnote 20. After removing duplicates in Endnote 20, the articles were exported to Covidence, a collaborative software. The search was closed on 30^th^ September 2024.

#### Study selection

Once the articles were imported into Covidence, duplicates were removed again. The review process then started. It consisted of two levels of screening: (1) a title and abstract review and (2) full-text review. GM and a co-reviewer (ON) did the first and second level of screening. The inclusion and exclusion criteria specified the following:

1. Articles must be published between 1^st^ January 2000 and 30^th^ September 2024.
2. Articles must be in English.
3. Articles must report findings from primary research.
4. Articles must fit within the PCC framework.

Any articles that did not fit these criteria were excluded. The articles that had a title and abstract that seemed to be eligible were included in this first level of screening. These articles reported on FSWs and had some aspects of participation in SHG within the SSA context.

The second stage of the review involved GM and ON reading through all the articles that were included for the full-text review. To determine inter-rater agreement for articles to include in full text review, Covidence software was used. Any discordant full-text articles were reviewed a second time and further disagreements about study eligibility at the full-text review stage were resolved through discussion with a more senior researcher (WM) until full consensus was obtained.

## Results

### Study descriptions

The search yielded 1,567 potentially relevant records from the three databases; 92 records were identified by Covidence to be duplicates and 1,474 records remained for title and abstract screening. Title and abstract screening were done by two reviewers and there were 36 conflicts from the records (i.e., reviewers disagreed). The conflicts were mainly on articles that were not SHG-related but seemed to look at issues to do with empowerment. The review team resolved conflicts by going through the titles and abstracts together. There remained 35 articles for full text screening. After cross-validation and full text screening, 11 articles were selected. Studies that were: not from SSA, did not focus on FSWs, were not within the study period and, did not employ primary research methodologies were excluded. Figure 1 below presents a summary of the selection process.

**Figure 1:**
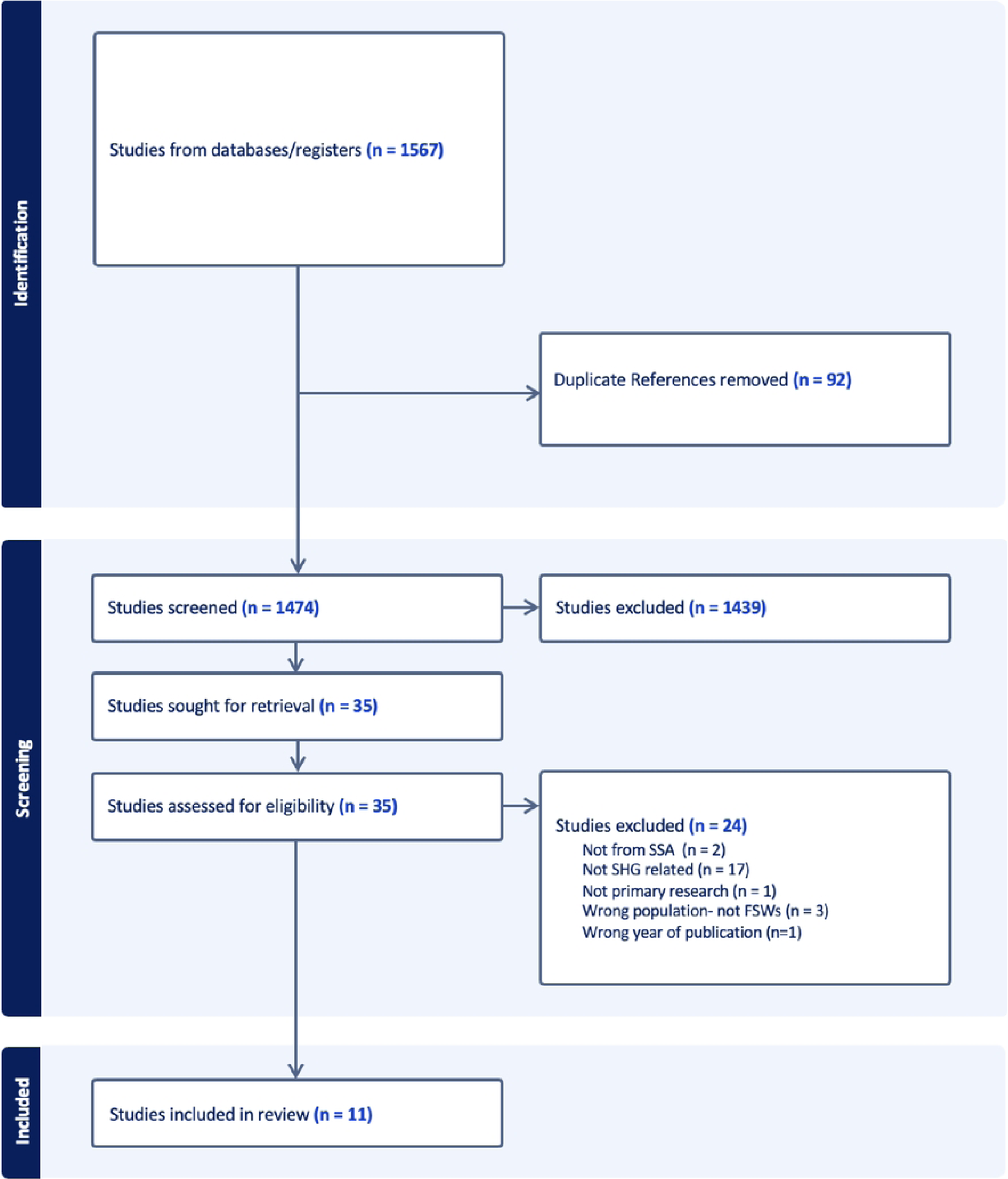
PRISMA flowchart of study selection process.

### Study characteristics

The 11 studies included for final review were conducted in seven countries in SSA. One study was multi-country and conducted in three countries: Kenya, Tanzania, and Uganda [23]. Four of the studies were conducted in Tanzania and were of the same intervention and trial (Project Shikamana, 4.1 to 4.3) [24–27]. One study was conducted in Zimbabwe [28], another was from Cameroon [29], one from Côte d’Ivoire [30], one from South Africa [31] and two from Kenya [32, 33] (Table 2).

**Table 2:**
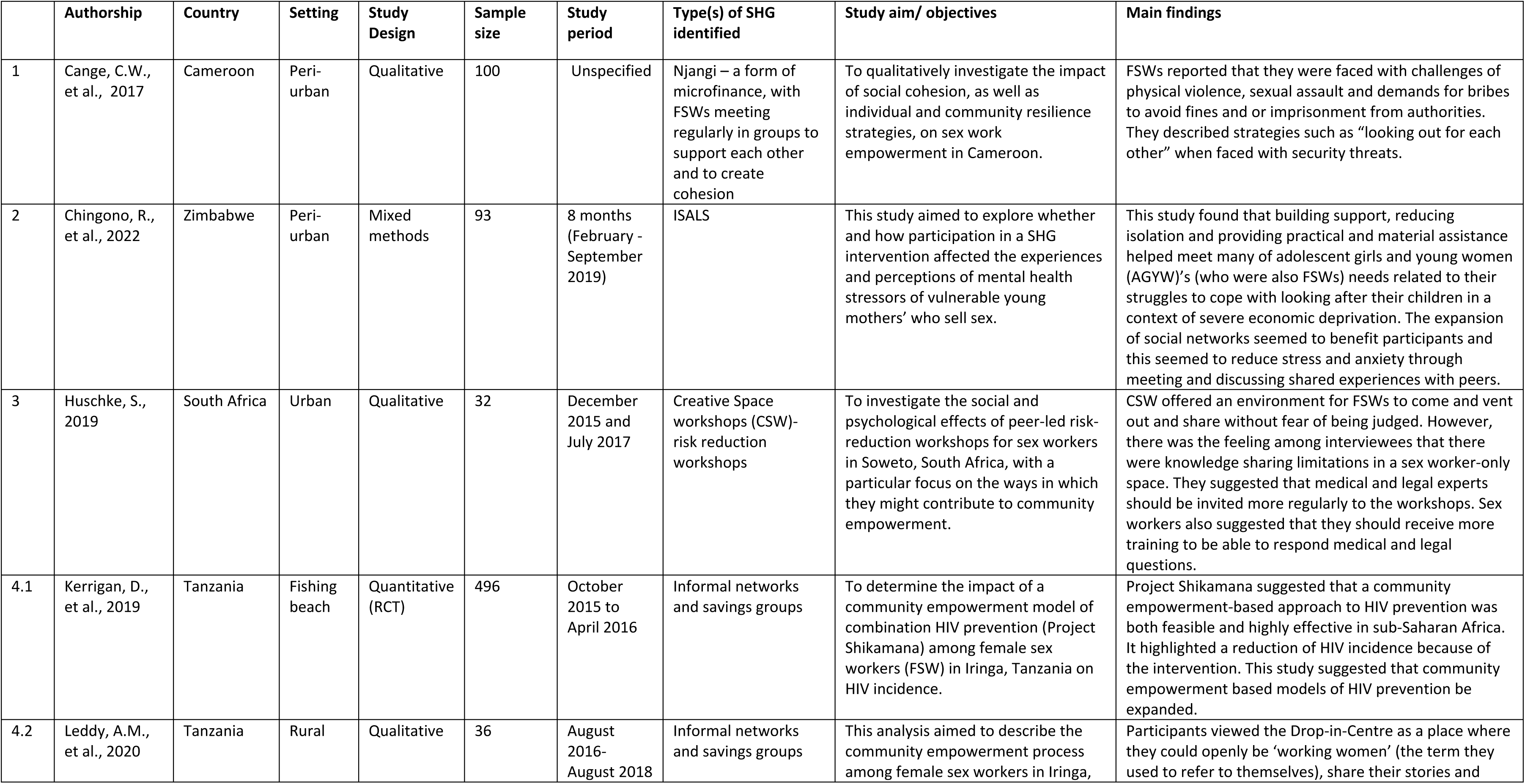
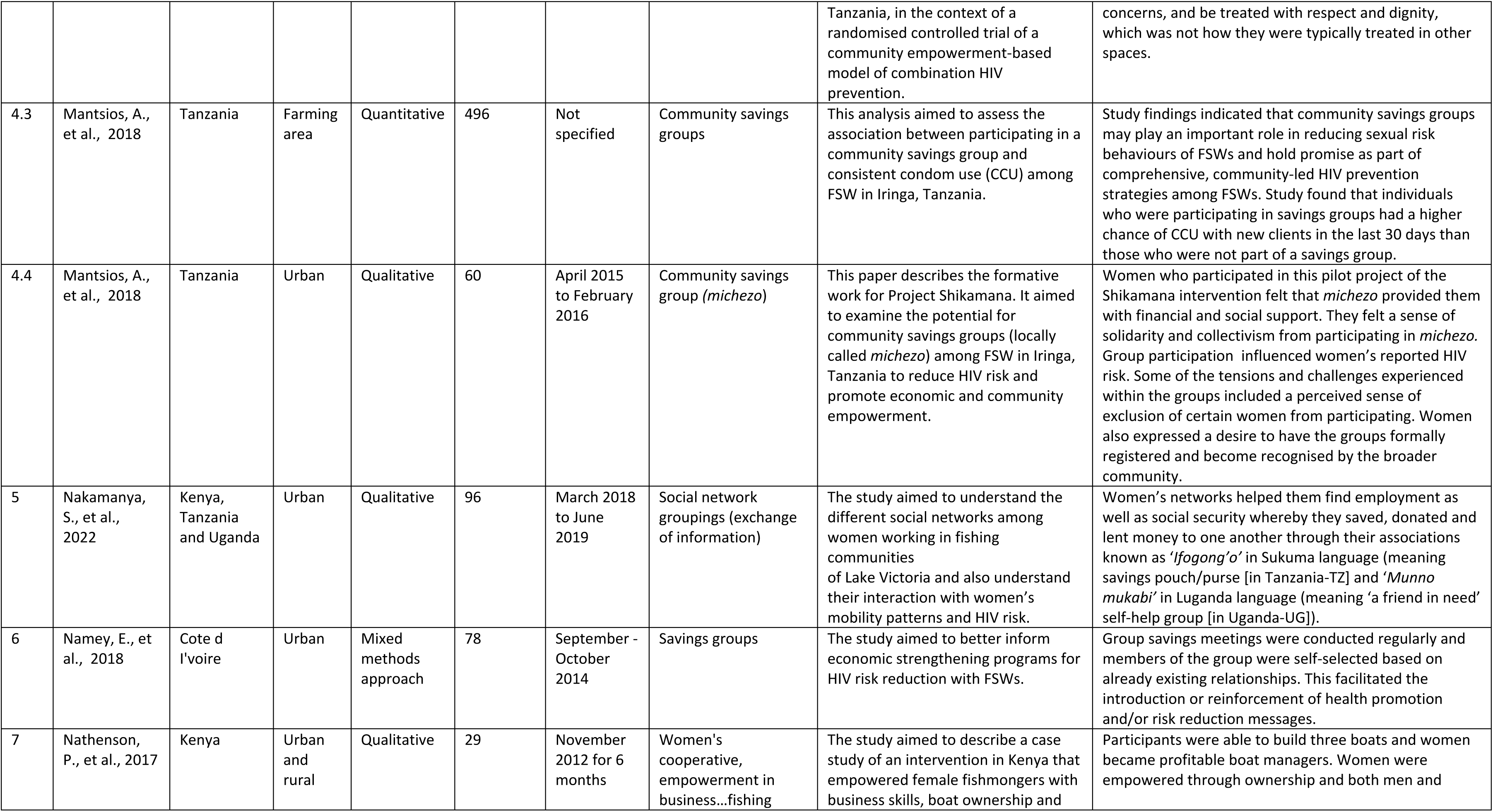
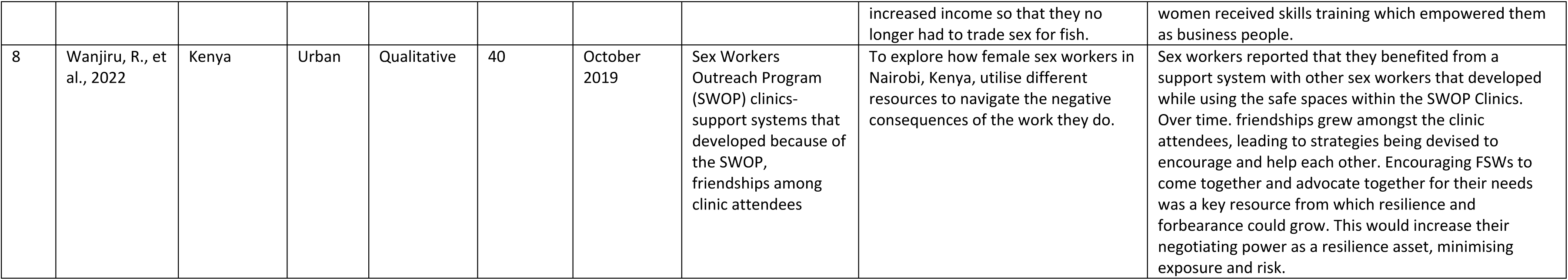
Study characteristics.

While the review primarily focused on FSWs, two of the articles were diverse and FSWs were studied within the context of other populations such as fishermen, HIV care providers, police, venue managers, community advisory board members and research staff [23, 34] (Table 2).

### Quality of included studies

The quality of included qualitative studies was assessed using the Critical Appraisal Skills Programme (CASP) tool (Table 3) [35], which comprises ten questions. Of the nine qualitative studies included, one was a case study and could not be assessed using all the ten questions. We determined that three of the ten CASP questions were not relevant to the case study. All the included studies were deemed to be of good quality based on the answers to the ten CASP questions.

**Table 3:** Critical Appraisal Skills Programme (CASP) Qualitative Research Checklist (2022)

|  | Section A: Are the results valid? |  |  |  |  |  | Section B: What are the results? |  |  | Section C: Will the results help locally? |
| --- | --- | --- | --- | --- | --- | --- | --- | --- | --- | --- |
| Publication | Q1: Was there a clear statement of the aims? | Q2: Is a qualitative methodology appropriate? | Q3: Was the research design appropriate to address the aims of the research? | Q4: Was the recruitment strategy appropriate to the aims of the research? | Q5: Was the data collected in a way that addressed the research issue? | Q6: Has the relationship between researcher and participants been adequately considered? | Q7: Have ethical issues been taken into consideration? | Q8: Was the data analysis sufficiently rigorous? | Q9: Is there a clear statement of findings? | Q10: How valuable is the research? |
| Cange, C.W., et al., 2017 | Yes- the study sought to qualitatively investigate the impact of social cohesion, as well as individual and community resilience strategies, on sex work empowerment in Cameroon | Yes | Yes: a qualitative research design was appropriate to respond to the aims of exploring the impact of social cohesion | A qualitative research design was appropriate to respond to the aims of exploring the impact of social cohesion.<br>In as much as the recruitment strategy was mentioned initially, it is not clear how the 37 participants eventually made it to the final sample. | Yes: the use of in-depth interviews and focus groups was in my view adequate to obtain data that spoke to the research question | Yes... interviews were conducted in a language well understood by participants and researchers were trained to speak to sex workers.<br>However, there is no statement of researcher positionality. | The study obtained requisite IRB clearance as well as written consent from participants | Yes- poor and inconsistent quality transcripts were eliminated and there was inter-researcher triangulation. | Yes- 4 key themes were derived from the findings | The research is valuable in showing how valuable social cohesion is especially when they come together in groups to address common challenges; e.g. going to the clinic together, loaning each other money in groups. However, this study did not go into much detail about the organisation of these groups that the FSWs were in and it lacks enough evidence on effectiveness of SHGs in empowering FSWs. |
| Chingono, R., et al., 2022<br>(mixed methods) | Yes- to examine participants' mental health stressors and how these may have been influenced through participation in the SHG intervention. | Yes- Using FGDs and semi structured interviews as well as questionnaires | Yes, the research design was adequate to address the aims of the research | Yes | Yes, data was adequately selected | Not really:<br>The lead author does mention her positionality as a female social scientist, but the paper does not address the researcher-participants relationship in clearer detail. However, in terms of the data collection in the methods section, there is an indication that the relationship between the researcher and the participants was taken into consideration. | Yes, ethical clearance was obtained from regulatory authorities. Also, written informed consent from participants | Yes, the data analysis was rigorous enough as the authors explained how they formulated the content analysis and subsequent inductive coding framework | Yes, four key themes were derived from the findings | The research is valuable as it shows the effectiveness of SHGs in impacting mental health among young FSWs. It also highlights why some SHG members drop out and this feeds into the question on sustainability |
| Huschke, S., 2019 | Yes- The aim of this paper is to investigate the social and psychological effects of Creative Space workshops for sex workers in Soweto, South Africa, with a particular focus on the ways in which they might contribute to community empowerment. | Yes, a qualitative design was appropriate since the study sought to explore the efficacy of peer-led workshops in empowering FSW | Yes: The research design was relevant to address the aims of the study | The paper doesn't specify how the participants were recruited. It however mentions that the data that was analysed was a subset of data that was collected earlier in an ethnographic study. | Yes. Data collection spoke sufficiently to the research issue | The relationship between the researcher and the participants has not been adequately considered. However, there is mention that interviews were conducted in Isizulu, but it is not explicitly mentioned that this was the language of the participants. I am drawn to assume that this was the language of the participants | Yes- the study received ethical approval. However, consent was oral and not documented | Data analysis was rigorous, but the main shortcoming was that it was done by one person. It is not mentioned in the paper whether the author was working with other researchers to triangulate themes coming from the analysis of the data and there is risk of subjectivity. | Yes- findings are very clear and supported by quotes. | The research is very valuable as it espouses how female sex workers coming together in groups can facilitate the process of empowerment as they support each other to navigate through common challenges through the CSWs. |
| Leddy, A.M., et al., 2020 | Yes, it aimed to describe the community empowerment process among female sex workers in Iringa, Tanzania, in the context of a randomised controlled trial of a community empowerment-based model of combination HIV prevention. | Yes- A qualitative approach was ideal to respond to the aims of the study | Yes- The research design was necessary to address the study aims | Yes- Recruitment was correct as it drew participants from those initially selected for the RCT | Yes- Use of in-depth interviews and key informant interviews to gather qualitative data | Is not stated, however, its indicated that the interviews were conducted in the language of the participants | Yes, there was ethical approval from IRB | Yes- The study adopted a deductive-inductive thematic analysis approach | Yes- Yes, the findings are clearly stated and logically presented | The results will help in giving insights on how SHGs can be better sustained and develop to be more structured. |
| Mantsios, A., et al., 2018 | Yes, examines the potential for community savings groups (locally called *michezo*) among FSW in Iringa, Tanzania to reduce HIV risk and promote economic and community empowerment. | Yes, a qualitative methodology was followed and adequately spoke to the aims of the study | Yes, the research design was qualitative and spoke to the aims of the study | Yes- Recruitment was correct as snowball sampling was used to gather the sample. | Yes- Qualitative data collection tools were used while nonprobability sampling methods were also used. | Yes- interviewer was trained and had established rapport with the participants. Interviews were also conducted in the local language | Yes- Participants provided oral consent and their identities were anonymised. Relevant institutional permissions to conduct the study were sought and granted | Yes- Data were analysed manually as well as through qualitative data management and analysis software (ATLAS Ti) | Yes- Findings were thematically presented and supported with quotes | The results are very useful in understanding savings groups among FSWs. It clearly defined how the group was organised and how it operated and this speaks to sustainability of such groups. |
| Nakamanya, S., et al., 2022 | Yes- to describe different social networks, mobility patterns and risk of HIV acquisition among women resident and/or working in fishing communities in the three East African countries bordering Lake Victoria: Kenya, Tanzania and Uganda | Yes- A qualitative approach is warranted in this study | Yes- the multi-site qualitative nature of the study is explicitly stated in the paper | Yes- recruitment was guided by the gatekeepers across the 4 sites. However there is likely to have been bias as they may have subjectively influenced choice of respondents | Yes- Yes, data were collected qualitatively through participant observation, and in depth interviews | Yes- interviews conducted in the languages of participants and conducted by trained interviewers | Yes- Community entry and Informed consent were sought from community elders/gatekeepers and participants respectively | Yes-thematic framework approach was used and the process was iterative | No- There is no clear statement of findings, however, different findings are presented in the results section | The results will help in showing the downside of groups which come together and actually live together and move together in cases of migration. The findings also help look at FSWs in the context of other population groups that they work alongside; e.g. fishermen, bar workers etc. |
| Namey, E., et al., 2018<br>(mixed methods) | Yes-to help describe potential distal financial determinants of HIV risk and to inform the development of a pilot economic strengthening program. | Yes- A mixed methods approach informed this paper | Yes- Yes, the approach was relatively appropriate | Yes- Yes, the recruitment spoke to the aims of the study | Yes- Mixed data were collected with qualitative data and quantitative financial data being collected | Case workers from local organisations who identified the participants for the study were the ones who introduced participants to interviewers. However, no other information shows the relationship between the participants and the interviewers<br>Selection of study participants may have been influenced by the recruitment methods of using case workers... there may be subjectivity | No- Participants granted verbal/oral informed consent | Yes, data was analysed using thematic analysis | Yes- Data were presented following specified thematic codes | This study did not give more insights about SHGs. The focus was more on how FSWs manage their finances rather than on SHGs. It lack enough evidence on SHGs and how they influence FSWs. |
| Nathenson, P., et al., 2017 | Yes- This paper describes a case study of an intervention in Kenya that empowered female fishmongers with business skills, boat ownership and increased income so that they no longer had to trade sex for fish. | Yes | Not clear- the methodology section is unclear, but as a case study, it managed to address the aim of the study | Methodology section unclear | Unclear | Unclear | Unclear | Unclear | Clear outcomes were documented | Results will be useful locally as it clearly shows the impact of self-help groups through empowerment initiatives (savings fund) |
| Wanjiru, R., et al., 2022 | Yes- to explore how different experiences over the life-course of FSWs interact to keep them disenfranchised or marginalized, especially in Kenya and explore how they cope, survive, and even thrive, despite these circumstances. | Yes- it is appropriate as the paper seeks to explore resilience issues | Yes-it was appropriate as it used the in-depth interview method to collect data | Yes- Participants were randomly sampled | Yes- Data were collected through IDIs | Yes- the social scientist who were trained extensively on the protocol and they interviewed ion the language of the participants | Yes | Yes, three themes identified | Yes | Yes-information on how they built resilience through their support systems informs how SHGs can facilitate empowerment of FSWs |

To assess the quality of the one randomised control trial (RCT) that was included, we used the Cochrane Risk of Bias 2 (RoB 2) tool [36] (Table 4). This tool assesses five aspects of bias: selection, performance, detection, and reporting. Risk categories were very low, low, moderate, and high.

**Table 4:** Quality appraisal for RCT (Risk of bias 2 (RoB2)

|  | Domain | Risk of bias | Risk of bias judgement |
| --- | --- | --- | --- |
| 1 | **Selection bias** | The study used a 2-community randomised trial design, which reduces the risk of selection bias. Time-location sampling was used to enrol participants, which is a robust method for reaching hard-to-reach populations like female sex workers. The baseline characteristics of the intervention and control groups appear to be well-balanced, further reducing the risk of selection bias. | Very low |
| 2 | **Performance bias** | The study description indicates that the intervention involved multiple components, including a community-led drop-in centre, peer education, service navigation, and provider trainings. These intervention elements were delivered to the intervention group, while the control group did not receive these activities. This differential delivery of the intervention creates a potential risk of performance bias. | Low |
| 3 | **Detection bias** | The study reports that HIV testing and viral load assessments were conducted at baseline and 18-month follow-up. These outcome assessments were done using standard laboratory methods, which reduces the risk of detection bias. However, the document does not provide details on whether the outcome assessors were blinded to the participants' intervention status. | Low |
| 4 | **Attrition bias** | The study had some loss to follow-up, with 171 HIV-positive and 216 HIV-negative participants completing both baseline and 18-month assessments. The authors used intent-to-treat analysis and inverse probability weighting to account for missing data, which helps mitigate the risk of attrition bias. | Low |
| 5 | **Reporting bias** | The study appears to have reported on all the pre-specified primary and secondary outcomes, reducing the risk of reporting bias. The protocol or trial registration is not provided, so there is some uncertainty about potential selective reporting. | Low |
| 6 | **Other bias** | The study was funded by the US National Institute of Mental Health, and there is no indication of other sources of funding that could introduce bias. The authors declare no conflicts of interest, further reducing the risk of other biases. | Low |
|  | **Overall** | Overall, this was a well-designed and conducted study with a generally **low** risk of bias, though some uncertainty remains around blinding of outcome assessors and potential selective reporting. | Low |

One included cross-sectional study was assessed using the Risk Of Bias In Non-randomised Studies of Interventions (ROBINS-I) assessment tool [37] (Table 5). This tool focuses on bias: due to confounding, in study participant selection, in interventions classification, due to deviations from intended interventions, due to missing data, in outcome measurement and in selection of the reported result. Risk categories were low, moderate, and high.

**Table 5:** Critical appraisal of cross-sectional quantitative study-Risk Of Bias In Non-randomised Studies of Interventions (ROBINS-I)

|  | **Domain** | **Explanation** | **Risk of bias judgement** |
| --- | --- | --- | --- |
| 1 | **Bias due to confounding** | The study adjusted for potential confounders like age, education, and marital status in the multivariable analysis. However, there may be other important confounders, such as duration of sex work, that were not adjusted for. | Moderate |
| 2 | **Bias in selection of participants into the study** | Bias in selection of participants into the study was low because time location sampling was used to recruit study participants. | Low |
| 3 | **Bias in classification of interventions** | In this observational study, the key "intervention" was participation in community savings groups. The study states that a validated measure was used to assess this exposure. | Low |
| 4 | **Bias due to deviations from intended interventions** | Not directly applicable. | - |
| 5 | **Bias due to missing data** | The study used complete case analysis, indicating minimal missing data on the key variables. | Low |
| 6 | **Bias in measurement of outcomes** | The primary outcome of consistent condom use was self-reported by participants. While validated measures were used, self-reported sexual behaviours are subject to social desirability bias. | Moderate |
| 7 | **Bias in selection of the reported result** | The study appears to have reported all the pre-specified analyses without evidence of selective reporting. | Low |
|  | **Overall risk of bias assessment** | Based on the ROBINS-I assessment, the overall risk of bias for this cross-sectional observational study is **low**. The main sources of potential bias are in the areas of confounding and the use of self-reported outcome measures. |  |

Overall, all the included studies were of good quality.

## Discussion

Various theories have been used to explain how SHGs can influence health outcomes of women and these span various fields. We adapted the integrated empowerment framework for FSWs which combines various elements and strategies aimed at empowering individuals or communities. The three main domains of this framework are power within, power with (others), and power over (resources) [13, 38, 39]. We borrow from some components of the framework [13]. The empowerment framework is depicted in Figure 2 below.

**Figure 2:**
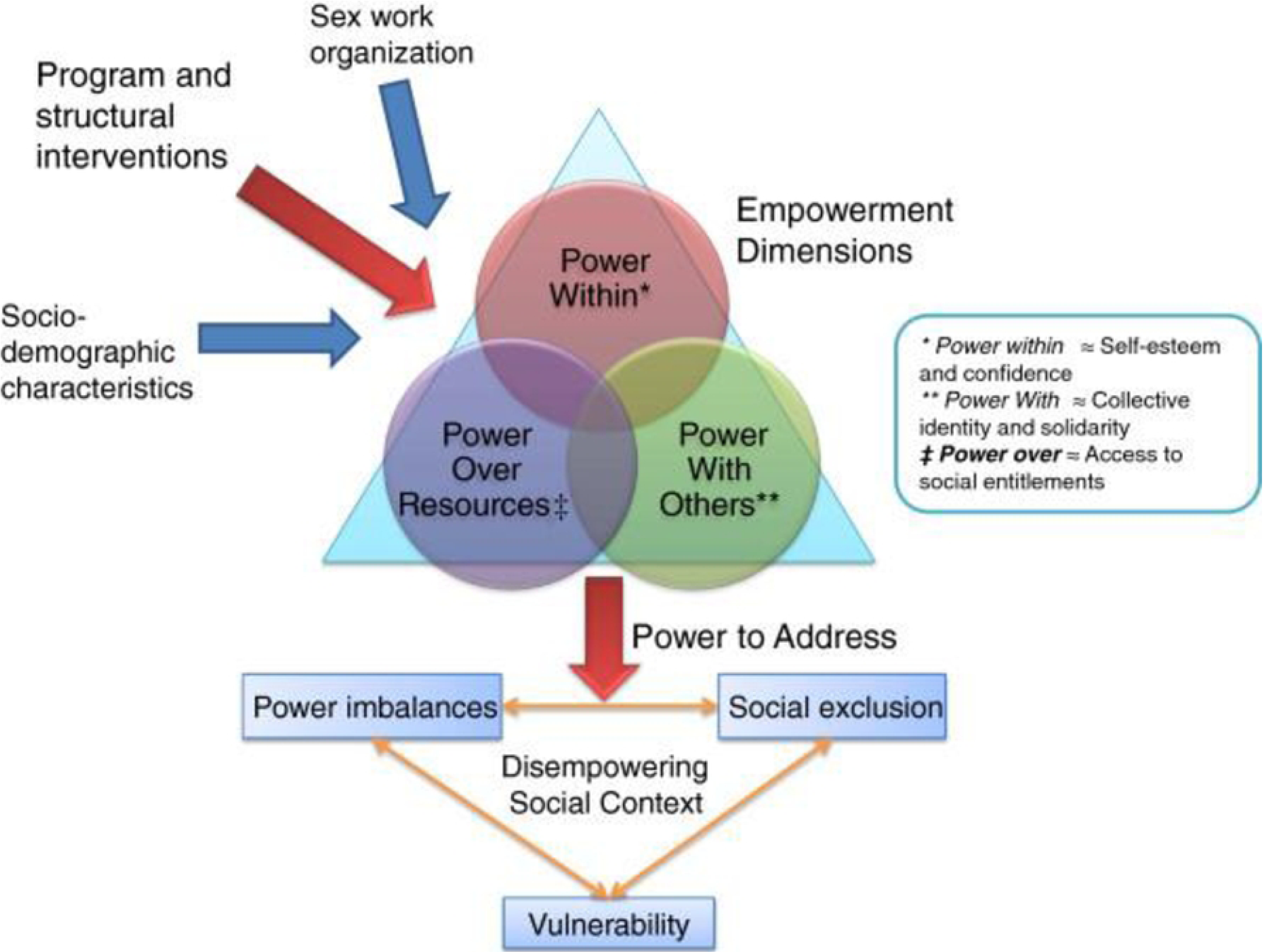
Integrated empowerment framework. Source: Blanchard et al. BMC Public Health 2013, 13:234

### Power within

The concept of “power within” refers to the extent of influence, control, and decision-making authority that individuals or groups possess within their immediate social and environmental contexts [40]. It relates to the degree to which an individual or group can exercise their agency and affect their own lives or the lives of others within their microsystem [40].

Being a member of a SHG empowered FSWs with knowledge of HIV prevention services. For example, in the Kenyan study, most of the participants mentioned having little or no knowledge of HIV prevention before engaging with the Sex Workers Outreach Program (SWOP) clinic services [30, 32]. However, women reported that after accessing the services and information from the SWOP’s ‘sex worker-friendly’ clinics, they no longer agreed to have sex without a condom because they understood that the nature of the relationship with their clients was purely transactional [32]. In the SWOP clinics, some participants also reported using pre-exposure prophylaxis because they were now more aware of the risks involved, and should there be a condom failure, they were protected [30]. The empowerment they gained by being part of a SHG helped FSWs recognise their self-worth and choose whom to have sex with, and reduced their reliance on men [30]. This Kenyan study suggests that other health outcomes, such as modern family planning use, may also be influenced by these interventions, especially when family planning is part of a comprehensive empowerment-based program [32].

Two studies found statistically significant associations between higher pay per sexual encounter, higher total income, higher sex work income and group participation [24, 26]. Community savings group participation was significantly associated with reported consistent condom use (CCU) with new clients and regular clients (but not with steady, non-paying partners) [26]. Higher income was associated with CCU with a new partner[26]. CCU with a regular partner was also associated with higher income and other factors such as savings group participation, older age, longer time in sex work, and having financial dependents. Overall, SHGs enhanced FSWs’ agency, which aligns with the “power within” concept.

### Power with others

“Power with others” denotes a collaborative and shared form of authority where individuals or groups come together in pursuit of common objectives or collectively address issues [40]. It embodies the concept that power is not concentrated within a single entity or individual; instead, it is distributed among the participants in a cooperative manner [40]. Within this framework, individuals or groups join forces, combining their collective resources, knowledge and influence so that they effect change, make decisions, or achieve tasks, usually to aid fairness, social justice, or mutual benefit[40].

Most of the FSWs reported benefiting from a support system with other sex workers, their local community, that they had devised to encourage and help each other out [28, 32]. This support included, but was not limited to, financial and emotional support, sharing of health education information and linkage to health care services [32]. Others also highlighted how they helped each other to seek medical assistance during illness and, in some cases, even accompanying each other to the clinic/hospital for treatment [27, 28, 41]. Where they faced violence from clients, they assisted each other through telephone-based security strategies where they called each other for assistance [42]. Additionally, they offered each other guidance on issues such as condom breakage, partner violence and matters related to child support and care. It is also within these SHGs that they advised each other on the dangers of drug and alcohol use. These bonds and associations within their peer groups provided solace and the means to thrive despite numerous challenges [32].

Social capital (i.e. benefits derived from association) was a resource for resilience which held great value among FSWs. Furthermore, the act of coming together allowed FSWs to address instances of abuse by security forces during their work [42]. FSWs also trained each other on how to survive in the industry and on social security. Also, they taught each other about condom use [23, 24, 27].

Creative Space workshops (CSWs) offered FSWs a platform to address common shared vulnerabilities. FSWs recognised the therapeutic value of peer-led group sessions, where they shared their personal struggles and traumas. These sessions contributed to enhancing sex workers’ self-esteem by fostering a supportive, non-judgmental, rights-based discourse, which led to a reframing of sex work as legitimate employment and portrayed sex workers as individuals with inherent rights [31]. They also exposed sex workers to valuable health knowledge, broadened their understanding of their legal rights, and informed them on available support services. In this way, CSWs promoted a greater sense of emotional well-being, and was experienced as empowering, instilling a greater sense of agency in the face of pervasive structural and interpersonal challenges. It also fostered hope that their circumstances could improve. Overall, the group concept and collective empowerment illustrated the “power with others” concept.

### Power over resources

“Power over resources” pertains to the capacity of individuals, organisations, or entities to exercise control, distribute, and make decisions regarding assets, commodities, or properties [43]. This power capacitates them to determine how these resources are used, distributed, and managed [43]. It is an essential aspect of resource management and can have significant implications for individuals and communities and how they manage SRH and vulnerability to HIV [43].

In the context of savings funds, that is, where FSWs jointly contribute some money and the disbursement of funds within SHGs, they were generally perceived as empowering [41], and empowerment has a positive influence on sexual risk behaviour. Although some SHGs gave FSWs power over resources, they fell short of establishing a framework for FSWs to secure and manage long-term savings. These savings would typically be earmarked for purposes such as transitioning from sex work, homeownership, child support, marriage, accessing essential healthcare, or returning to their places of origin [44]. However, one study clearly demonstrated the “power over resources” when FSWs were able to register a catering company business [25]. They formalised their activities by registering with the government as an official business and this business grew, registering membership of 50 FSWs and became self-sustained and continued meeting even after the trial [25]. However, it was not established in this study how far this empowerment impacted on SRH and HIV outcomes.

The benefits derived from participating in SHGs for FSWs predominantly revolve around the empowerment of the individual (power within) and the strength they gain from uniting as a group (power with others). However, in most of the articles reviewed, achieving “power over resources” remains a challenge. Empowerment strategies have been employed to increase FSWs’ access to social entitlements, financial credit, and educational opportunities which opens up more choices for them in terms of decisions that impact their HIV and SRH outcomes. In India, for example, significant progress has been made with regards to achieving “power over resources” but this is something that takes time [45]. One good example is the Usha Multi-purpose Cooperative Society Limited (USHA), which is the largest and first ever sex worker-led financial institution in Southern Asia which is exclusively run by sex workers and for sex workers [45]. Since there is a need for comprehensive change, it is acknowledged that community mobilisation strategies among FSWs must be complemented by structural interventions aimed at addressing the underlying social, economic, legal, and political structures that initially contributed to their disempowerment.

Most of the included studies were of short duration, with most lasting less than one year. This relatively short intervention period may have limited the ability to observe the long-term results that are anticipated from participation in SHGs, including improved mental and physical health, reduced risk of HIV acquisition, and decreased HIV transmission, among other SRH concerns. Therefore, some of these desired long-term outcomes may not have been apparent due to the brief nature of the studies.

While numerous studies have underscored the manifold benefits of SHG participation, it is essential to recognise that they do not create an idealised, conflict-free environment where the challenges sex workers face in their daily lives cease to exist. One prominent issue highlighted in the literature pertains to the tensions that can arise between different groups of sex workers and between sex workers and non-sex workers who occasionally engage in these groups to access associated benefits [23, 31]. Trust emerges as another crucial factor that significantly influences the success of SHGs. In certain studies, trust issues were evident as some FSWs expressed reservations about trusting their peers, which, in turn, had repercussions for SHGs’ overall effectiveness.

### Strengths and limitations

This scoping review allowed us to broadly examine the different types of SHGs existing in SSA and assess how they influence SRH and HIV among FSWs. The focus on urban, rural, peri-urban and fishing communities allow for the data to be generalisable among broader population contexts. However, a few limitations can be noted. Firstly, we used only three search engines–Medline, Global Health and CINAHL–to conduct the article search despite knowing that other search engines may have yielded studies not included here. This was because conducting a scoping review involves substantial time and resources and inherent constraints led us to focus on the three well-curated and widely used databases. We carefully selected these three databases to minimise redundancy in our search results, considering the potential for overlapping content across multiple databases. They are comprehensive and they maximised the depth and relevance of the literature that was retrieved. This contributed positively to the overall quality of the review. Another limitation relates to the relatively few studies found, mostly published recently because SHGs are an emerging area of research in Africa, that needs to be expanded. Finally, we only included studies published in English which may have introduced language bias.

## Conclusions

SHGs have shown great promise in empowering FSWs and addressing their health vulnerabilities. To fully harness the potential of SHGs, it is crucial to adopt a holistic, long-term, and community-centred approach, address trust and conflict issues, and advocate for broader structural changes that support FSWs’ well-being and rights. By doing so, we can contribute to a more comprehensive and effective response to the complex health challenges faced by this vulnerable population.

## Data Availability

This scoping review synthesises existing literature on whether and how self-help groups improve female sex workers’ sexual and reproductive health and HIV outcomes in SSA. The data and studies included in this review are derived from publicly accessible sources. All articles reviewed are available through Medline, Global Health and CINAHL databases, and can be accessed without restriction. While the review does not present original data, it compiles findings from eleven studies published between 1 January 2000 and 30 September 2024. Interested researchers can access these studies directly through their respective journals or databases. The protocol for this study has been published and is accessible at the following DOI: [https://doi.org/10.12688/wellcomeopenres.23002.1]. This protocol outlines the methodology used in this study and is available for reference by researchers interested in replicating or building upon this work.

## Acknowledgments

We would like to acknowledge the support that was offered by the LSTM librarian who assisted in development of the search terms and manoeuvring the online library system.

## Supporting information

S1_Table: Mesh and free text search strategy

## Notes

### Competing Interest Statement

The authors have declared no competing interest.

### Funding Statement

This work was supported by Wellcome [214280]. The funders had no role in study design, data collection and analysis, decision to publish, or preparation of the manuscript. All arguments and opinions expressed in this protocol are solely of the authors.

### Author Declarations

As this study is a scoping review that synthesises existing literature and does not involve original data collection from human participants, ethical approval was not required. Therefore, there are no details of an IRB or oversight body to provide.

